# The effects of Autonomous Sensory Meridian Response (ASMR) on sleep quality improvement in adolescents

**DOI:** 10.1101/2024.09.14.24312582

**Authors:** Zuoda Wu, Chao He, Kai Zhao

## Abstract

**Background:** In recent years, there has been a notable surge in sleep-related challenges among adolescents. The emergence of Autonomous Sensory Meridian Response (ASMR) content across various social media platforms has sparked interest in its potential to address these issues. This study explored the therapeutic impact of ASMR on sleep problems in high school students.

**Methods:** The study involved sixty participants, divided evenly into four groups: three intervention groups (A, B, C) and one control group. For five consecutive days, the intervention groups engaged in pre-sleep ASMR listening sessions lasting 10, 20, and 30 minutes, respectively. Sleep quality was evaluated using mobile application tools and the subjective Sleep Quality Scale.

**Results:** The analysis, performed utilizing repeated measures sphericity tests, revealed a significant correlation between ASMR intervention days and both objective sleep(*P* = 0.036) and effective sleep duration (*P* = 0.002). However, ASMR intervention did not significantly affect objective sleep efficiency or subjective sleep quality. These findings imply that ASMR interventions can significantly improve sleep issues in adolescents.

**Conclusions:** ASMR has been extensively promoted as a potential remedy for sleep enhancement, this study substantiates its effectiveness in tackling adolescent sleep concerns.

## Introduction

Numerous studies indicate that a significant portion of Chinese high school students are grappling with increasingly severe sleep disturbances amidst escalating academic pressure and competition. (Chen et al. 2005; Gui et al. 2022; Xiong et al. 2023; Zhou et al. 2023). A meta-analysis of 66 studies revealed that 37.6% of Chinese children have sleep problems, with 11.3% experiencing restlessness and 11.1% struggling to fall asleep. Furthermore, the prevalence of sleep issues increased with age(Chen et al. 2021). Additionally, studies indicate that high school adolescents exhibit a higher prevalence of sleep problems compared to their junior high school counterparts (Liang et al. 2021).

The high school period is an important stage for the physical development and character formation of adolescents(Gautam et al. 2021; Huang et al. 2023). Sleep deprivation will adversely impact various aspects including obesity, cognitive development and depression(Zhai et al. 2018; Zhang et al. 2022; Zhao et al. 2022). Hence, it is imperative to allocate adequate attention and implement effective interventions to enhance the sleep quality of Chinese high school students.

In recent years, the concept of auditory intervention, particularly Autonomous Sensory Meridian Response (ASMR), has gained prominence. ASMR refers to a physiological sensation triggered by specific visual and auditory stimuli (Mahady et al. 2023). ASMR encompasses various elements including image guidance, progressive relaxation, hypnosis, meditation and other contents(Sakurai et al. 2021), and it can be used as a tool for enhancing sleep quality(Lee et al. 2019). ASMR, characterized by low-frequency human voice and ambient sounds, has garnered significant attention across social media platforms, short and long-form video platforms, and podcasts, owing to its unique pleasurable and stimulating effects(Ahuja & Ahuja 2019). By exploring the experiences of insomniacs, it is possible to design sleep-assisting applications centered around ASMR content.

Currently there are few studies on the relationship between sleep problems and ASMR in high school students, particularly in terms of quantifying this relationship(Liu & Zhou 2019). Therefore, this study will explore the feasibility of utilizing ASMR as an auditory intervention for addressing sleep problems among adolescents. The goal is to provide effective recommendations to ameliorate sleep disturbances among Chinese high school students.

## Methods

### 1. Recruitment of subject

Sixty volunteers (30 males and 30 females) aged 15-19 were recruited from six high schools in China. These participants had watched ASMR videos but had never felt the characteristic tingling sensation. Additionally, they had no psychiatric disorders and were not on any medication. To prevent habituation effects, the volunteers were prohibited from watching any ASMR content for one week before the experiment began. The experiment was approved by the Ethics Committee of Tongji Medical College, Huazhong University of Science and Technology.

### 2. ASMR selection

ASMR videos can be broadly divided into two categories: non-dramaturgic (e.g., trigger sounds, white noise, meditative relaxation) and dramaturgic (e.g., role play, casual conversation). For this experiment, we focused on the non-dramaturgic type, selecting five videos featuring trigger sounds based on their popularity with the general public.

### 3. ASMR Intervention

The participants were randomly assigned to four groups: Group A (control group), Group B (listened to ASMR for 10 minutes), Group C (listened to ASMR for 20 minutes), and Group D (listened to ASMR for 30 minutes). The aim was to compare the effects across these groups to determine the most effective ASMR duration for enhancing sleep quality.

### 4. Sleep quality assessment

To assess sleep quality, we employed the subjective Pittsburgh Sleep Quality Index, allowing participants to rate their sleep quality. In addition, a mobile phone app was used to objectively gather data on sleep patterns, including objective sleep duration, effective sleep time, and overall sleep quality. Participants completed the questionnaire, and researchers compiled the scores.

### 5. Statistics analysis

This section underwent statistical analysis using Repeated Measures ANOVA. To meet the prerequisites of this analysis, initial assessments of normality and sphericity were conducted. Q-Q plots were utilized to assess normality, and non-normally distributed data were transformed using a natural logarithm (ln) for further analysis. The sphericity test was conducted as follows: a P-value less than 0.05 indicated a failure to meet sphericity, necessitating corrections. If sphericity was not met and Mauchly’s W exceeded 0.75, the Huynh-Feldt (HF) correction was applied. Conversely, a Mauchly’s W value below 0.75 led to applying the Greenhouse-Geisser (GG) correction. When sphericity was confirmed (P &gt; 0.05), no corrections were made to the within-group effects analysis. For post hoc analyses, the Bonferroni method was used to adjust P-values, ensuring statistical rigor.

## Results

### 1. Effects of ASMR Intervention on Objective Duration of Sleep

Quantile-Quantile (Q-Q) plots showed that objective sleep duration data were not normally distributed (**Figure 1A**). However, after applying a natural logarithm transformation, the data achieved normal distribution(**Figure 1B**). The data failed the sphericity test (P = 0.046), and Mauchly’s W was under 0.75, leading to the selection of the Greenhouse-Geisser (GG) correction for further analysis in the repeated measures ANOVA (**Table 1**).

**Table 1.** Tests of Sphericity.

| Outcomes | Mauchly's W | P-value | Sphericity correction |
| --- | --- | --- | --- |
| Objective Time to Sleep | 0.729 | 0.046 | Greenhouse-Geisser |
| Effective Length of Sleep | 0.808 | 0.236 | / |
| Objective Sleep Efficiency | 0.198 | <0.001 | Greenhouse-Geisser |
| Subjective Sleep Quality | 0.600 | 0.001 | Greenhouse-Geisser |

**Figure 1.**
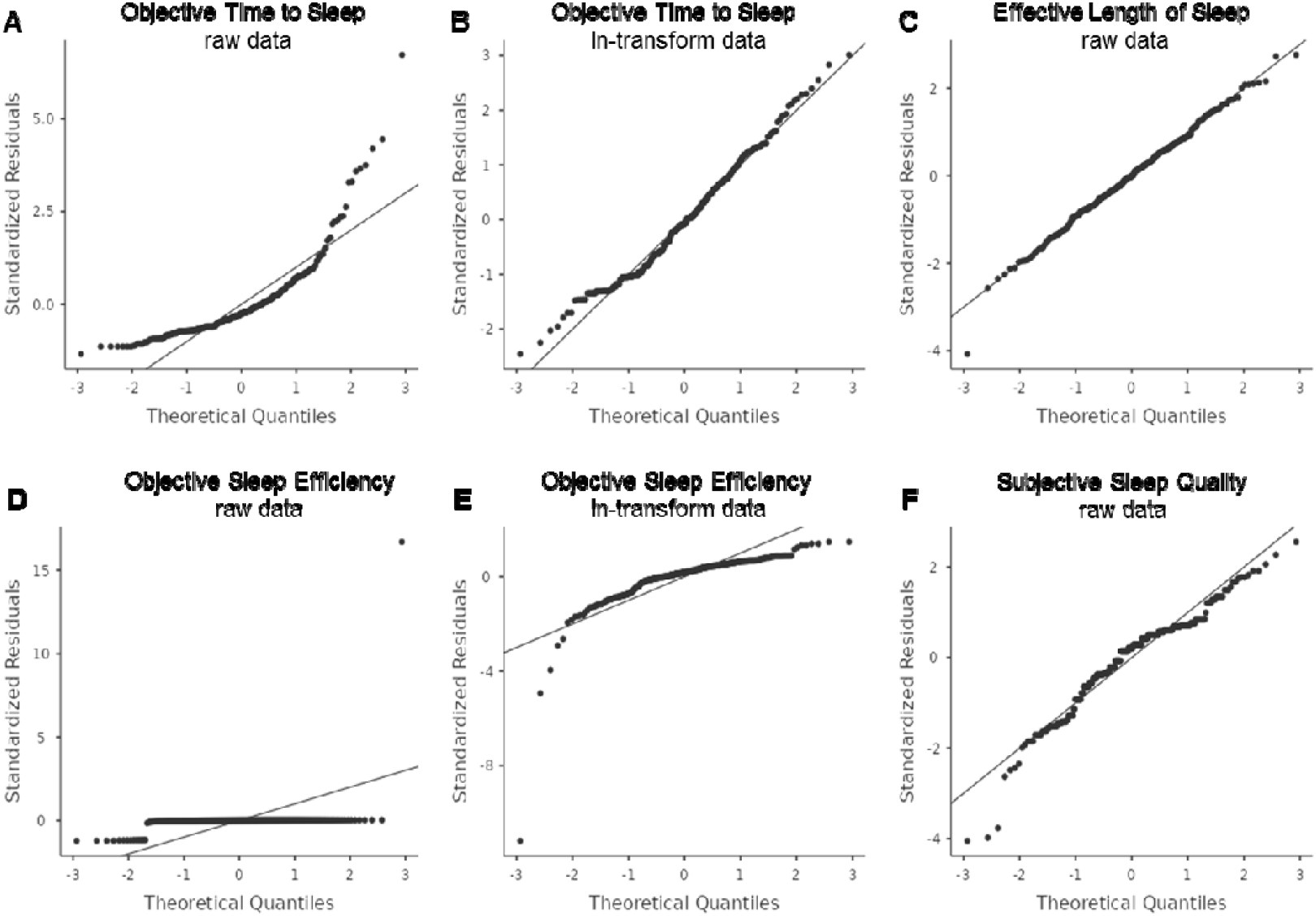
Q-Q plots of normality of observed data. A. The raw data of objective sleep duration; B. The natural log-transformed data of objective sleep duration; C. The raw data of effective sleep duration; D. The raw data of objective sleep efficiency; E. The natural log-transformed data of objective sleep efficiency; F. The raw data of subjective sleep quality score.

The within-group analysis of the repeated measures ANOVA identified a statistically significant variation in the onset of objective sleep across different ASMR intervention days (*P* = 0.036, η^2^ = 0.027), indicating that the number of days of ASMR intervention influenced the timing of sleep onset. However, no interaction was found between the number of ASMR intervention days and the duration of each session (*P* = 0.967, ^η2^ = 0.011) (**Table 2**).

**Table 2.** Within Subjects Effects.

| Outcomes | F | P-value | Effect Size |  |  |  |
| --- | --- | --- | --- | --- | --- | --- |
| | | | $\eta^2G$ | $\eta^2$ | $\eta^2p$ | $\omega^2$ |
| Objective Time to Sleep |  |  |  |  |  |  |
| Times | 2.753 | **0.036** | 0.029 | 0.027 | 0.047 | 0.017 |
| Times:Treatment | 0.355 | 0.967 | 0.011 | 0.011 | 0.019 | <0.001 |
| Effective Length of Sleep |  |  |  |  |  |  |
| Times | 4.332 | **0.002** | 0.051 | 0.046 | 0.072 | 0.035 |
| Times:Treatment | 0.742 | 0.709 | 0.027 | 0.024 | 0.038 | <0.001 |
| Objective Sleep Efficiency |  |  |  |  |  |  |
| Times | 0.550 | 0.591 | 0.007 | 0.007 | 0.010 | <0.001 |
| Times:Treatment | 1.024 | 0.415 | 0.038 | 0.038 | 0.052 | 0.001 |
| Subjective Sleep Quality |  |  |  |  |  |  |
| Times | 1.605 | 0.188 | 0.022 | 0.021 | 0.028 | 0.008 |
| Times:Treatment | 0.151 | 0.998 | 0.006 | 0.006 | 0.008 | <0.001 |

The between-group effects analysis revealed no significant impact of the intervention length on sleep onset (P = 0.112)(**Table 3**). Subsequent post hoc analysis also found no notable effect of ASMR intervention days on the length of objective sleep onset(**Table 4**). **Figure 2** shows the estimated marginal means for the ASMR intervention days and the duration of each intervention in relation to the length of objective sleep onset.

**Table 3.** Between Subjects Effects.

| Outcomes | F | P-value | Effect Size |  |  |  |
| --- | --- | --- | --- | --- | --- | --- |
| | | | $\eta^2G$ | $\eta^2$ | $\eta^2p$ | $\omega^2$ |
| Objective Time to Sleep | 2.090 | 0.112 | 0.043 | 0.041 | 0.101 | 0.021 |
| Effective Length of Sleep | 4.934 | **0.004** | 0.076 | 0.071 | 0.209 | 0.056 |
| Objective Sleep Efficiency | 0.434 | 0.730 | 0.006 | 0.006 | 0.023 | <0.001 |
| Subjective Sleep Quality | 2.787 | **0.049** | 0.034 | 0.033 | 0.132 | 0.021 |

**Table 4.** Post Hoc Comparisons – Times.

| Times1 | Times2 | Mean Difference | SE | t | P (Bonferroni) |
| --- | --- | --- | --- | --- | --- |
| Day.1 | Day.2 | 0.083 | 0.117 | 0.711 | >0.999 |
| Day.1 | Day.3 | 0.331 | 0.133 | 2.498 | 0.154 |
| Day.1 | Day.4 | 0.315 | 0.127 | 2.484 | 0.160 |
| Day.1 | Day.5 | 0.282 | 0.159 | 1.772 | 0.819 |
| Day.2 | Day.3 | 0.248 | 0.116 | 2.146 | 0.362 |
| Day.2 | Day.4 | 0.232 | 0.102 | 2.263 | 0.275 |
| Day.2 | Day.5 | 0.198 | 0.133 | 1.489 | >0.999 |
| Day.3 | Day.4 | -0.016 | 0.134 | -0.120 | >0.999 |
| Day.3 | Day.5 | -0.050 | 0.123 | -0.405 | >0.999 |
| Day.4 | Day.5 | -0.034 | 0.132 | -0.254 | >0.999 |

**Figure 2.**
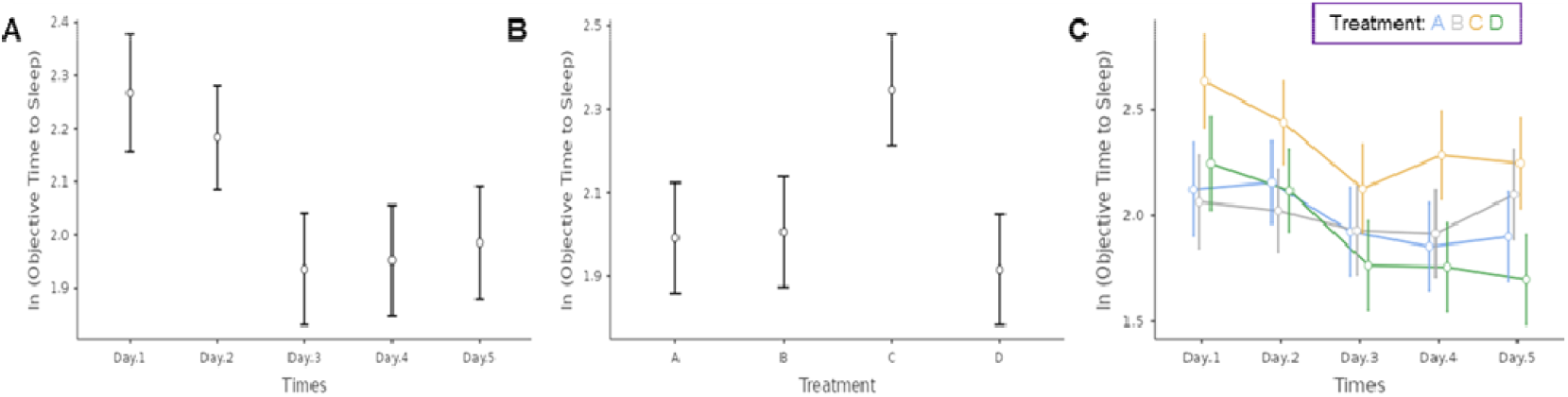
Comparison of subgroups of estimated marginal means of objective sleep duration. A. By ASMR intervention days; B. By ASMR intervention time per intervention; C. By ASMR intervention days and intervention time per intervention.

### 2. Effect of ASMR intervention on effective sleep duration

The Q-Q plot analysis revealed that the distribution of effective length of sleep hours was normal (**Figure 1C**) and met the sphericity criterion(*P* = 0.236), eliminating the need for adjustments in the subsequent repeated measures ANOVA(**Table 1**). The repeated-measures ANOVA within-group analysis indicated a significant difference in effective sleep duration across different ASMR intervention days (*P* = 0.002, ^η2^ = 0.046), suggesting a notable effect of the intervention frequency on sleep duration. However, no significant interaction was observed between the number of intervention days and the duration of each session (*P* = 0.709, ^η2^ = 0.024) (**Table 2**).

In the between-group effects analysis, the duration of ASMR interventions per session significantly impacted effective sleep duration(*P* = 0.004) (**Table 3**). In post hoc tests, compared with ASMR intervention day 1, ASMR intervention day 2 and day 5 were increased in effective sleep duration by 0.725 ± 0.229 (*P* = 0.025) and 0.943 ± 0.264 (*P* = 0.007), respectively (**Table 5**). Additionally, compared with Group B, interventions in Groups A and C resulted in significant increases in effective sleep duration of 0.914 ± 0.294 (*P* = 0.018) and 1.022 ± 0.294 (*P* = 0.006). **Figure 3** illustrates the estimated marginal means for the number of ASMR intervention days and session lengths in relation to effective sleep duration.

**Table 5.**
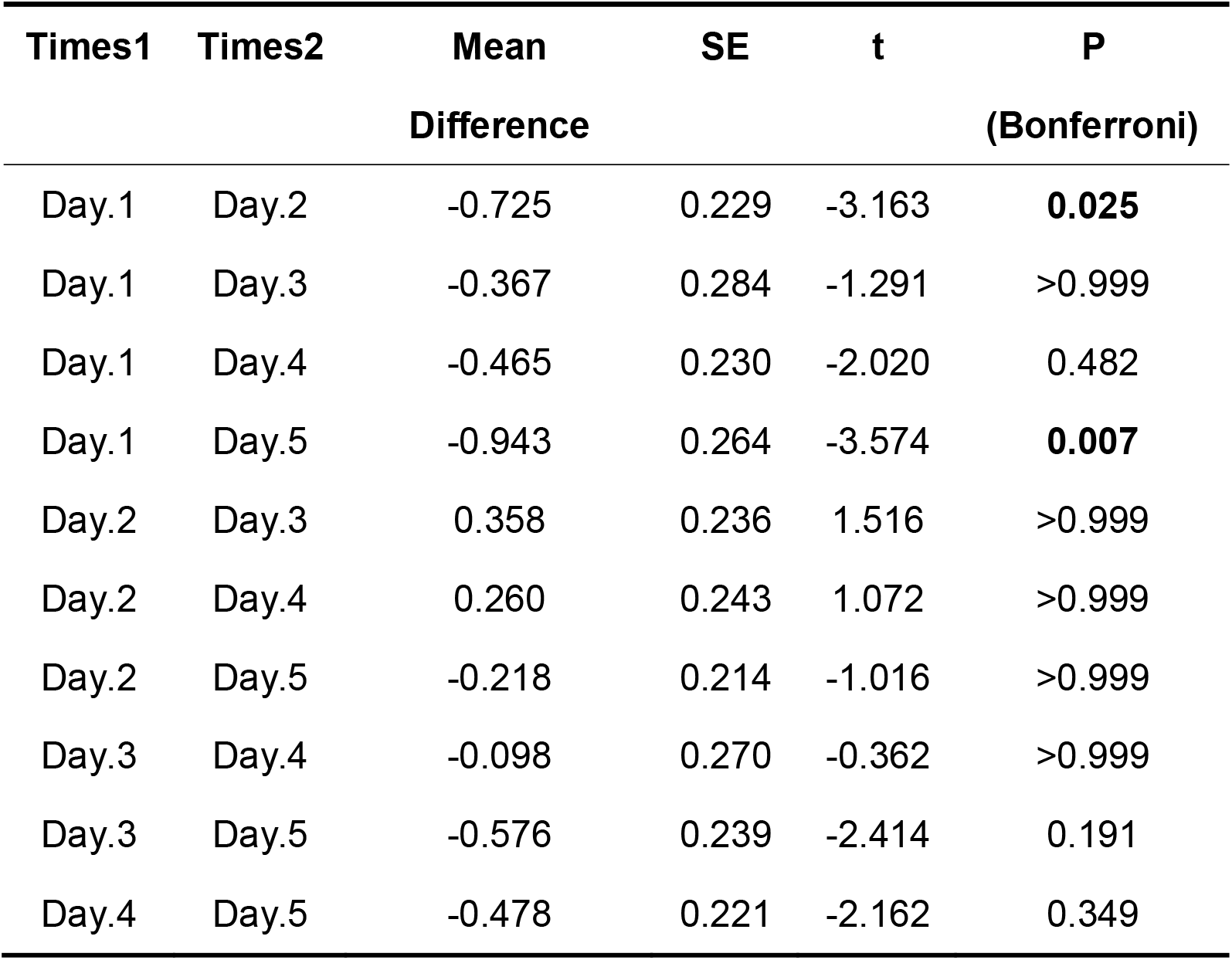
Post Hoc Comparisons – Times.

| **Times1** | **Times2** | **Mean** | **SE** | **t** | **P** |
| --- | --- | --- | --- | --- | --- |
|  |  | **Difference** |  |  | **(Bonferroni)** |
| Day.1 | Day.2 | -0.725 | 0.229 | -3.163 | **0.025** |
| Day.1 | Day.3 | -0.367 | 0.284 | -1.291 | >0.999 |
| Day.1 | Day.4 | -0.465 | 0.230 | -2.020 | 0.482 |
| Day.1 | Day.5 | -0.943 | 0.264 | -3.574 | **0.007** |
| Day.2 | Day.3 | 0.358 | 0.236 | 1.516 | >0.999 |
| Day.2 | Day.4 | 0.260 | 0.243 | 1.072 | >0.999 |
| Day.2 | Day.5 | -0.218 | 0.214 | -1.016 | >0.999 |
| Day.3 | Day.4 | -0.098 | 0.270 | -0.362 | >0.999 |
| Day.3 | Day.5 | -0.576 | 0.239 | -2.414 | 0.191 |
| Day.4 | Day.5 | -0.478 | 0.221 | -2.162 | 0.349 |

**Table 6.** Post Hoc Comparisons – Treatment.

| **Treatment** | **Treatment** | **Mean** | **SE** | **t** | **P** |
| --- | --- | --- | --- | --- | --- |
| **1** | **2** | **Difference** |  |  | **(Bonferroni)** |
| A | B | 0.914 | 0.294 | 3.113 | **0.018** |
| A | C | -0.108 | 0.294 | -0.369 | >0.999 |
| A | D | 0.367 | 0.294 | 1.251 | >0.999 |
| B | C | -1.022 | 0.294 | -3.482 | **0.006** |
| B | D | -0.547 | 0.294 | -1.862 | 0.407 |
| C | D | 0.476 | 0.294 | 1.620 | 0.665 |

**Table 7.** Post Hoc Comparisons – Treatment.

| **Treatment** | **Treatment** | **Mean** | **SE** | **t** | **P** |
| --- | --- | --- | --- | --- | --- |
| **1** | **2** | **Difference** |  |  | **(Bonferroni)** |
| A | B | 0.413 | 0.170 | 2.434 | 0.109 |
| A | C | 0.173 | 0.170 | 1.021 | >0.999 |
| A | D | 0.415 | 0.173 | 2.403 | 0.118 |
| B | C | -0.240 | 0.170 | -1.413 | 0.979 |
| B | D | 0.002 | 0.173 | 0.011 | >0.999 |
| C | D | 0.242 | 0.173 | 1.400 | >0.999 |

**Figure 3.**
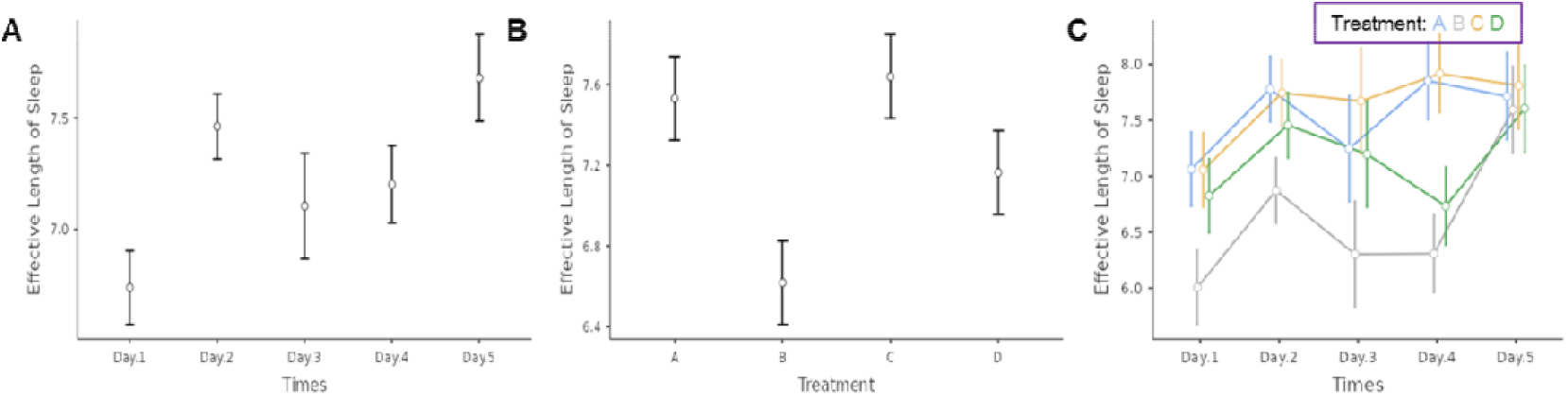
Comparison of subgroups of estimated marginal means of effective sleep duration. A. By ASMR intervention days; B. By ASMR intervention time per intervention; C. By ASMR intervention days and intervention time per intervention.

### 3. Effect of ASMR intervention on objective sleep efficiency

The analysis failed the sphericity test (P < 0.001) with Mauchly’s W falling below 0.75, leading to the selection of the GG correction for the subsequent repeated measures ANOVA (**Table 1**). The within-group analysis of the ANOVA revealed no significant differences in objective sleep efficiency across different ASMR intervention days (P = 0.591, ^η2^= 0.007), and no significant interaction was observed between the intervention frequency and duration (P = 0.415, ^η2^= 0.038) (**Table 2**). Additionally, the between-group analysis did not demonstrate any significant impact of the ASMR intervention duration on objective sleep efficiency (P = 0.730) (**Table 3**).

### 4. Effects of ASMR intervention on subjective sleep quality

Within-group results from the ANOVA indicated no significant variance in subjective sleep quality across different ASMR intervention days (P = 0.188, η^2^ = 0.021), and no interaction between the frequency of ASMR interventions and their respective durations (P = 0.998, η^2^ = 0.006) (**Table 2**). In contrast, the between-group analysis suggested that the length of each ASMR intervention influenced subjective sleep quality (P = 0.049) **(Table 3)**, although subsequent post hoc testing did not confirm a significant impact of ASMR duration per intervention on subjective sleep quality (**Table 4)**. Figure 4 presents the estimated marginal means of the ASMR intervention days and duration per intervention in relation to subjective sleep quality.

**Figure 4.**
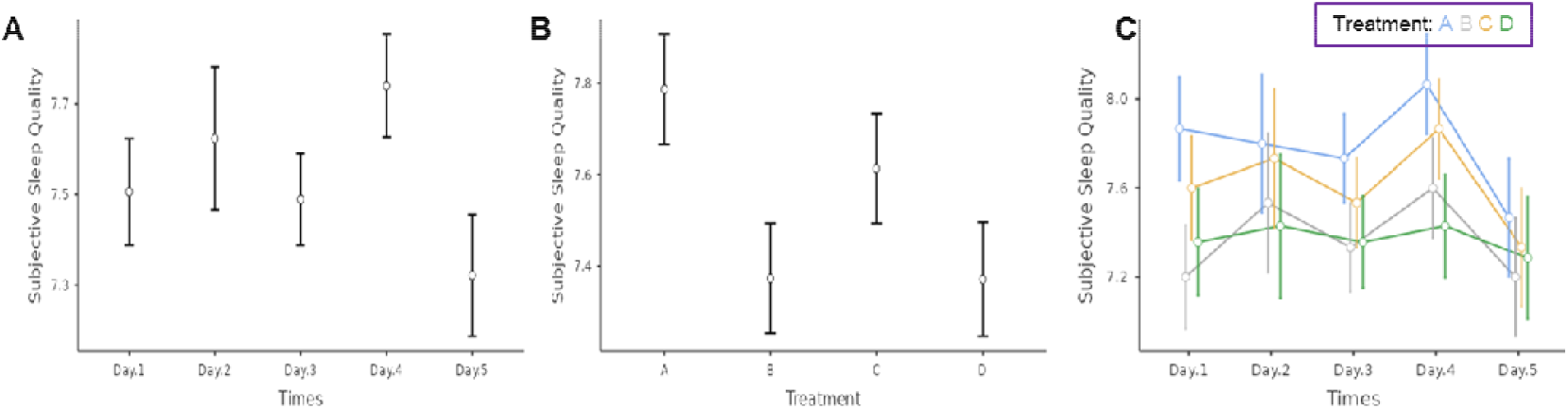
Subgroup comparisons of estimated marginal means of subjective sleep quality. A. By ASMR intervention days; B. By intervention time per ASMR intervention; C. By ASMR intervention days and intervention time per intervention.

## Discussion

In this study, we conducted a preliminary investigation into the impact of ASMR, a novel and popular tool, on the sleep quality of high school students. Our findings suggest a significant association between ASMR and improved sleep quality among high school students. These results offer insights into addressing high school students’ sleep issues through non-pharmacological approaches.

Sleep plays a crucial role in healthy development and is essential for both physical and mental well-being(Futenma et al. 2023; Tan et al. 2015). There have been many studies of adolescent sleep problems, addressing that shorter sleep duration is associated with poor physical and mental health in children and adolescents(Liu et al. 2016; Ou & Ma 2023; Xie et al. 2023; Ye et al. 2023). A systematic review explored the relationship between objective and subjective sleep durations and various health indicators in children and adolescents, and found that longer sleep duration was associated with reduced adiposity indicators, improved emotion regulation, enhanced academic performance, and better quality of life/well-being(Chaput et al. 2016). Consistent with the findings of the study through the sleep questionnaire(Lane et al. 2021), our study found that the high school students have poor subjective sleep quality.

The guidelines of insomnia treatment recommend prioritizing the use of non-pharmacological methods, such as psychotherapy(Riemann et al. 2017), but there is little mention of specific and feasible solutions for adolescents’ sleep problems(Takano et al. 2023). The auditory interventions are the mainstay of interventions for sleep disorders(Park et al. 2023). Auditory interventions, which use sound transmission, are a key approach for addressing sleep disorders and are commonly used among high school students(Engelbregt et al. 2022). This study aims to investigate the efficacy of auditory interventions for improving sleep in high school students.

Listening to ASMR audio recordings has been found to aid in calming the mind, promoting relaxation, alleviating pain, and enhancing sleep quality(Smith et al. 2017). In our study, we selected popular ASMR content among high school students, focusing on role-playing triggers known to resonate with adolescents, with an eye towards future applicability and promotion(Barratt et al. 2017; Smith et al. 2020). However, the effects of ASMR may vary from person to person(Fredborg et al. 2017). It’s important to note that individual responses to ASMR influenced by prior exposure and personal receptivity, which warrants further exploration.

To enhance the reliability of our findings, we employed a rigorous methodology involving multiple ASMR interventions and repeated measurements across several days, capturing variations for a comprehensive and objective data set(Arroyo & Zawadzki 2022; Hoang & Liang 2023). Our study utilized established sleep questionnaires and sleep tracking applications, which employ FDA-approved algorithms based on sound monitoring, to ensure the objectivity of sleep data(Al Mahmud et al. 2022).Interestingly, our findings indicated that the duration of ASMR exposure directly influence sleep length or quality. However, ASMR may initially facilitate sleep, prolonged exposure does not enhance its effectiveness and may potentially disrupt sleep quality. An automatic shutoff feature for ASMR upon sleep detection could potentially improve sleep quality.

This study also has some limitations, there’s a need for more precise objective measures, continuous improvement of algorithms, and larger participant groups for more conclusive results. Furthermore, the impact of ASMR on children’s brain functions remains underexplored and merits further investigation.

## Data Availability

All data produced in the present study are available upon reasonable request to the authors

## Author contribution

Zuoda Wu: Conceptualization, Methodology, Manuscript writing original draft and revision. Chao He: Conceptualization, Manuscript review and editing, Kai Zhao: Conceptualization, Manuscript writing and revision, Supervision.

